# Mapping The Global Landscape Of Chikungunya Rapid Diagnostic Tests: A Scoping Review

**DOI:** 10.1101/2022.01.28.22270018

**Authors:** José Moreira, Patrícia Brasil, Sabine Dittrich, André M Siqueira

## Abstract

**Background:** Chikungunya virus (CHIKV) is a re-emerging arboviral disease and represents a global health threat because of the unprecedented magnitude of its spread. Diagnostics strategies rely heavily on reverse transcriptase-polymerase chain reaction (RT-PCR) and antibody detection by Enzyme-linked Immunosorbent assay (ELISA). Rapid Diagnostic Tests (RDTs) are available and promise to decentralize testing and increase availability at lower healthcare system levels.

**Objectives:** We aim to identify the extent of research on CHIKV RDTs, map the global availability of CHIKV RDTs, and evaluate the accuracy of CHIKV RDTs for the diagnosis of CHIKV.

**Eligibility criteria:** We included studies reporting symptomatic individuals suspected of CHIKV, tested with CHIKV RDTs, against the comparator being a validated laboratory-based RT-PCR or ELISA assay. The primary outcome was the accuracy of the CHIKV RDT when compared with reference assays.

**Sources of evidence:** Medline, EMBASE, and Scopus were searched from inception to October 13^th^, 2021. National regulatory agencies (European Medicines Agency, U.S. Food and Drug Administration, and the Brazilian National Health Surveillance Agency) were also searched for registered CHIKV RDTs.

**Results:** Eighteen studies were included and corresponded to 3722 samples tested with RDTs between 2005-2019. The most development stage of CHIKV RDTs studies was phase I (7/18 studies) and II (7/18 studies). No studies were in phase IV. The countries that manufacturer the most CHIKV RDTs were Brazil (*n*=17), followed by the USA (*n*=7), and India (*n*=6). Neither at EMA nor FDA, registered products were found. Conversely, the ANVISA has approved 23 CHIKV RDTs. Antibody RDTs (*n*=43) predominated and demonstrated sensitivity between 20% and 100%. The sensitivity of the antigen RDTs ranged from 33.3 to 100%.

**Conclusions:** The landscape of CHIKV RDTs is fragmented and needs coordinated efforts to ensure that patients in CHIKV-endemic areas have access to appropriate RDTs. Further research is crucial to determine the impact of such tests on integrated fever case management and prescription practices for acute febrile patients.

## Introduction

Chikungunya -a re-emerging arboviral disease caused by Chikungunya virus (CHIKV)-is transmitted by mosquitoes of the Aedes species, specifically *A. aegypti, A. albopictus*, and *A. polynesiensis* [1]. The disease is characterized by the classic triad of debilitating polyarthralgia, high-grade fever, and myalgia [1]. During the past years, we have seen an unprecedented magnitude of the disease spreading across the globe (i.e., 106 countries/territories reported autochthonous or travel-related transmission), affecting millions of people in the Americas, Asia, the Indian subcontinent, Europe, and in the Pacific islands [2].

One of the challenges imposed by CHIKV has been the correct identification of suspected individuals in the context of co-circulation of other arboviruses that present similarly in tropical regions [3]. Laboratory diagnosis has been mainly focused on either RNA or virus-specific antibody detection through RT-PCR and ELISA technique, respectively. However, such diagnostic technologies require complex instrumentation and are not easy to perform outside sophisticated laboratories in urban settings where trained personnel are available. Therefore, these tests are not accessible or affordable to patients at the lower healthcare system levels, where most CHIKV outbreaks occur. In contrast, rapid diagnostic tests (RDTs) promise to overcome some of these challenges by bridging many gaps along the diagnostic test pathway in CHIKV-endemic areas.

RDTs have become available for detecting CHIKV and are reported to have variable performance and operational characteristics [4–6]. Much remains unknown regarding how these tests increase the efficiency of the health systems if introduced appropriately, how acceptable they are for patients and health care providers, and how cost-effective they are, given the poor state of many countries’ economies primarily impacted by CHIKV. Thus, we aim to (i) identify the extent of research on CHIKV RDTs; (ii) provide a comprehensive landscape of CHIKV RDTs available globally; (iii) evaluate the performance of CHIKV RDTs for the diagnosis of CHIKV in symptomatic individuals when compared with a reference standard, and (iv) identify knowledge gaps and further research related to CHIKV RDTs.

## Methods

We followed the PRISMA Extension for Scoping Reviews (Prisma-ScR) guidance from the EQUATOR (Enhancing the QUAlity and Transparency Of health Research) Network [7]. The Prisma-ScR checklist is available in the Supplementary material.

### Eligibility criteria

Search terms were based on a PICO (population, intervention, comparator, and outcome) framework. The population encompassed symptomatic febrile individuals suspected of CHIKV infection. The intervention used CHIKV RDTs, either in developmental or commercially available, to diagnose CHIKV infection, with the comparator being a validated laboratory-based RT-PCR or ELISA assay. The primary outcome was the accuracy of the CHIKV RDT when compared with reference assays.

Articles were excluded if (i) the studies were reviews, case reports, or opinion articles; (ii) the studies evaluated the performance of reverse transcription loop-mediated isothermal amplification (RT-LAMP) assays; (iii) the studies were related to an outbreak investigation without the evaluation of the accuracy of CHIKV RDTs; (iv) the studies used an inappropriate study population (asymptomatic individuals); (v) the studies described inappropriate reference assays to assign true positive/true negative status to study samples; and (vi) studies that were related to other arboviruses.

### Operational definitions

1. CHIKV RDT was defined as a rapid (≤ 60 min) point-of-care (POC) assay that requires minimal instrumentation to provide actionable results.
2. We classified the stage of CHIKV RDT assay development in four phases: phase I, which consist of the prototype evaluation process; phase II evaluation under ideal conditions using convenience or archived samples; phase III evaluations under ideal conditions assessing the performance and operation characteristics of the index test in a target population; and phase IV which are assessments of the impact of diagnostics on the prevalence of infection, the incidence of infection, or incidence of complications.

### Information sources

Medline, EMBASE, and Scopus electronic databases were searched from inception to 13 October 2021 to identify relevant publications in peer-reviewed journals as original scientific research. Additional studies were identified through manual searches of the reference lists of identified papers. The electronic database search was supplemented by searching at major tropical medicine conference abstracts repositories and the manufacturer’s official website to seek relevant unpublished reports. The final search results were exported into Mendeley to manage citations identified.

In order to provide a comprehensive assessment of diagnostic products that are in the developmental phase and commercialization, we conducted searches in national regulatory agencies (i.e., European Medicines Agency, U.S. Food and Drug Administration, and the Brazilian National Health Surveillance Agency) websites looking for registered CHIKV RDTs and a free search through the Google search engine.

### Search

The search in Medline was performed using the following terms: (chikungunya OR “chikungunya virus” OR “chikungunya fever”) AND (“rapid diagnostic test” OR “rapid test”). There was no language or time restriction. After deleting duplicates, the literature review group systematically screened the title, abstract and full text of each study’s inclusion and exclusion criteria.

### Data charting process

Data were extracted independently from the selected studies by two authors and recorded into a standard form designed for this study. Discrepancies were resolved by mediation and discussion with other reviewers if needed. The standardized data abstraction tool captured the relevant information on key study characteristics and detailed information on all metrics used to estimate the accuracy of the CHIKV RDTs. Key variables that were systematically extracted include the year of investigation, geographical location, study design, type of RDT assay, time of illness onset to testing, reference assay, sample size, and diagnostic accuracy parameters (if available). If a study evaluated more than one RDT assay, we extracted the data related to each assay type. When articles did not provide sufficient information on relevant data, we contacted the authors via email for additional information.

### Critical appraisal of individual sources of evidence

The quality of each diagnostic accuracy study was assessed following QUADAS-2 guidelines [8].

### Synthesis of results

Data from all studies were aggregated, and frequency statistics were run to describe the population across all studies. Tableau Desktop Professional Edition (Tableau software, LLC, version 2021.1.0, Seattle, Washington, US) and GraphPad Prism (GraphPad Software, version 8.0, San Diego, California, U.S.) were used to represent the evidence visually.

## Results

### Search results

The initial search identified 271 potential studies for evaluation (Supplementary Figure 1). After duplicates were removed, a total of 185 citations were identified from searches of electronic databases. Based on the tile and the abstract, 96 were excluded, with 89 full-text articles retrieved and assessed for eligibility. The remaining 18 studies were considered eligible for this review (all apart from one reported diagnostic accuracy metric).

### Description of studies

A summary of the included studies is shown in Table 1. The main countries where the CHIKV patients were sourced were India (3/18 studies, 16.6%), Thailand (3/18 studies, 16.6%), Indonesia (2/18 studies, 11.1%), and Aruba (2/18 studies, 11.1%) [Supplementary Figure 2]. CHIKV RDTs studies were phase I (7/18 studies, 38.8%) and II (7/18 studies, 38.8%) in most included studies. Three studies were phase III [4,9]. No study was phase IV. Sample recruitment used case-control methodologies (13/18 studies,72.2%), a prospective cohort design (4/18 studies, 22.2%), or described the development of a pilot RDT assay (1/18 studies, 5.5%)[10]. Description of the tested population and the setting where they were applied was almost absent in the studies.

**Table 1.**
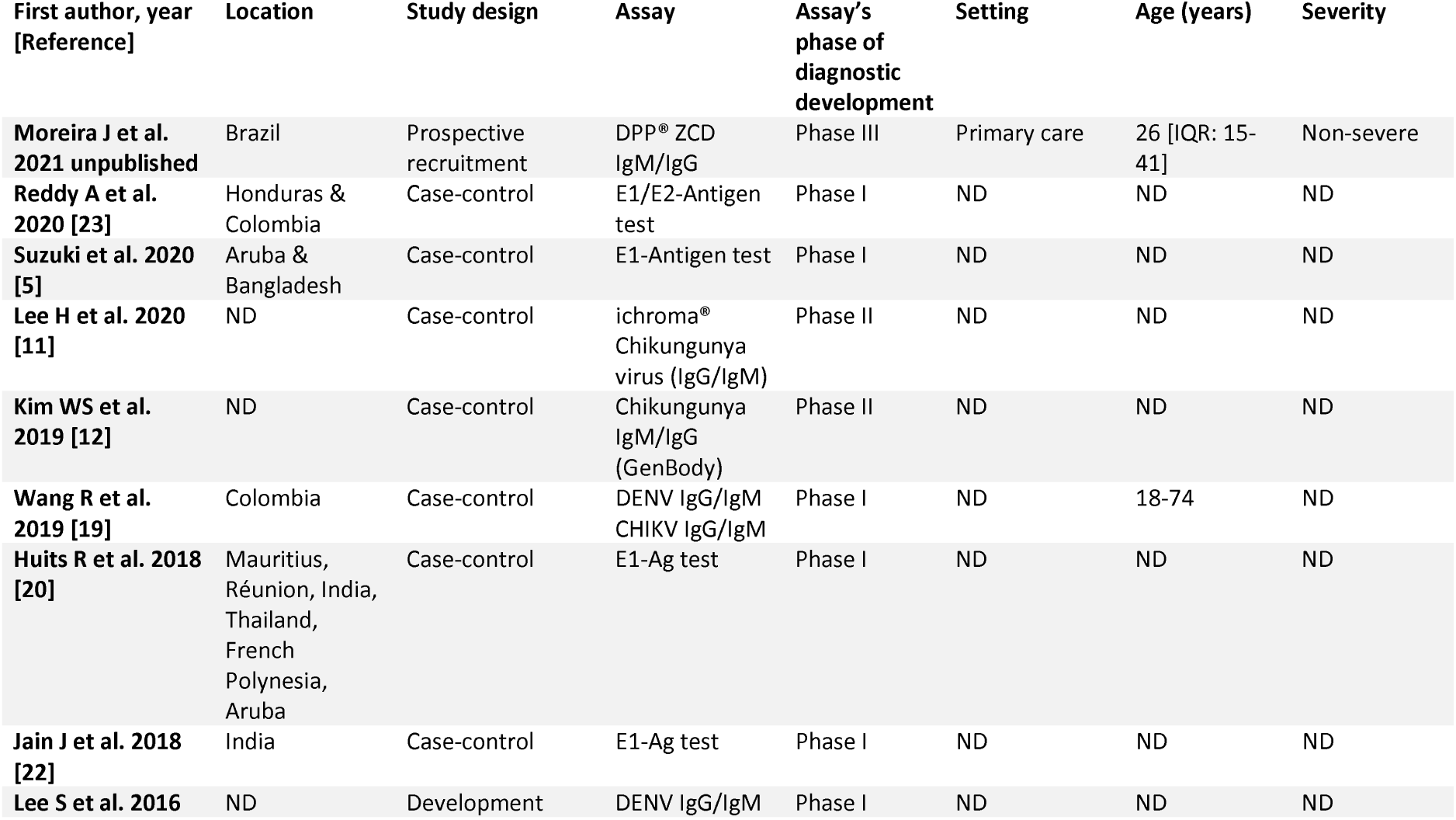
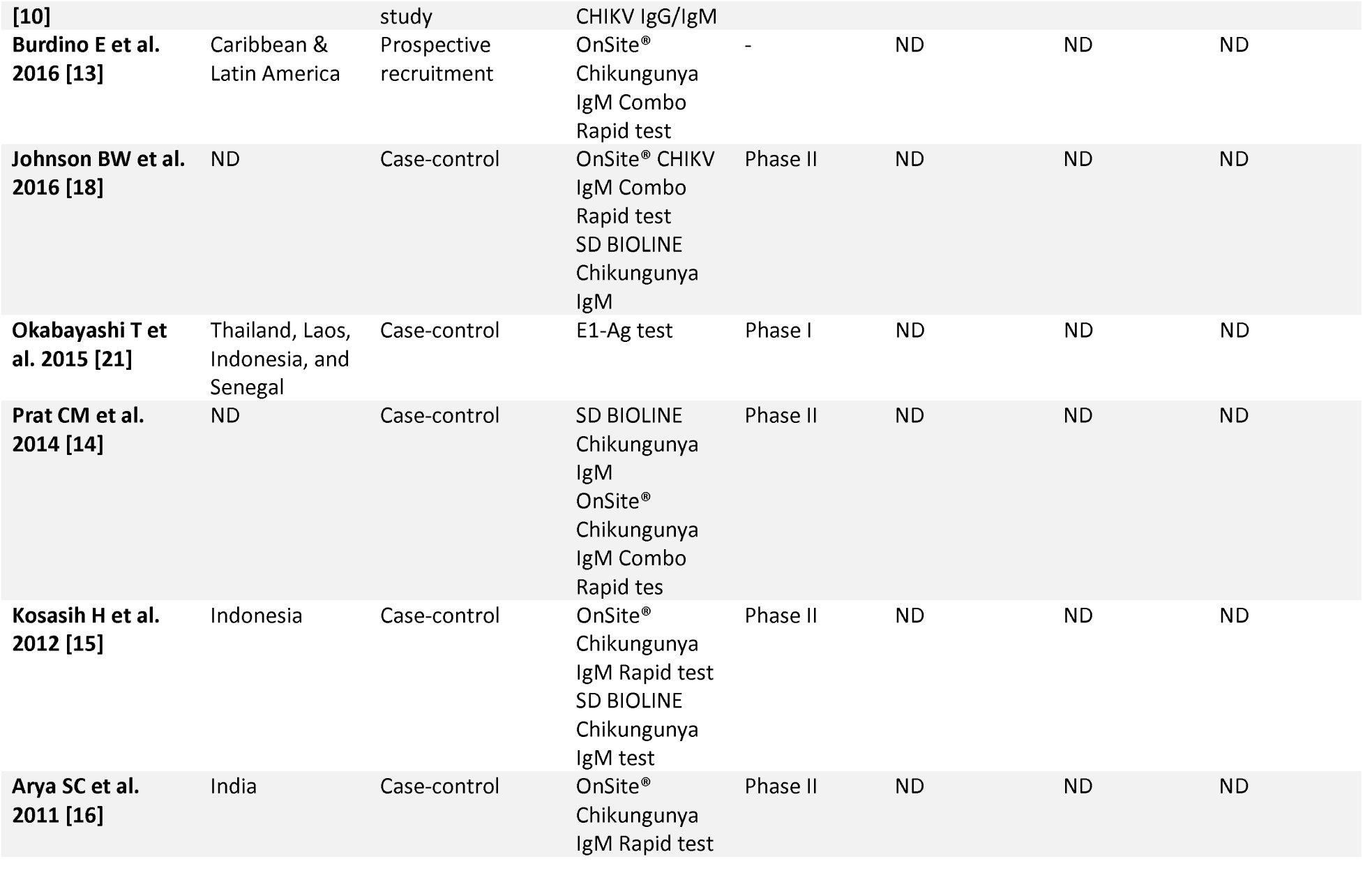
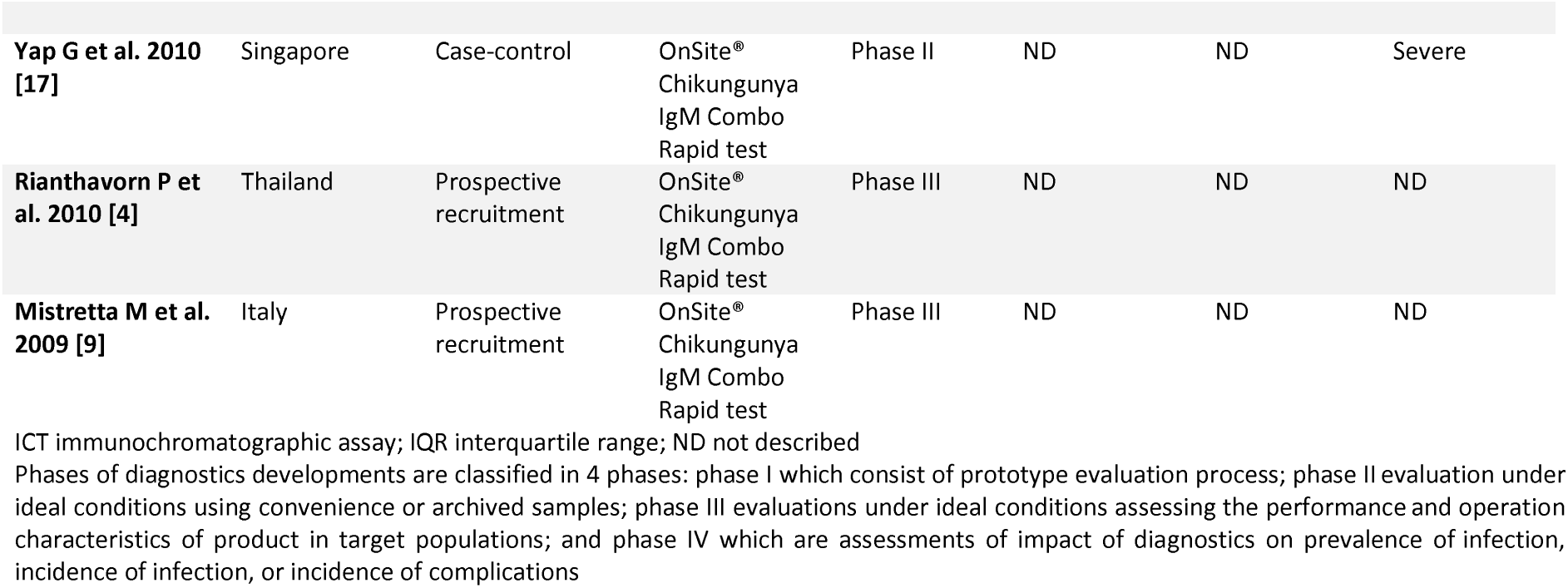
Characteristics of included studies evaluating Chikungunya antibody or antigen-based rapid diagnostic tests, 2005-2019.

### Global availability of Chikungunya RDTs

Table 2 shows the characteristics of CHIKV RDTs developed or commercialized for POC applications. The countries that manufacturer the most CHIKV RDTs were Brazil (*n*=17), followed by the USA (*n*=7), South Korea (*n*=7), and India (*n*=6) [Figure 1].

**Table 2.**
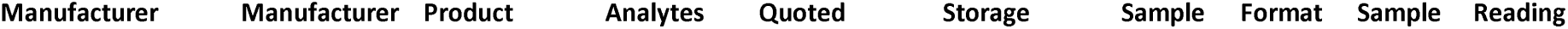
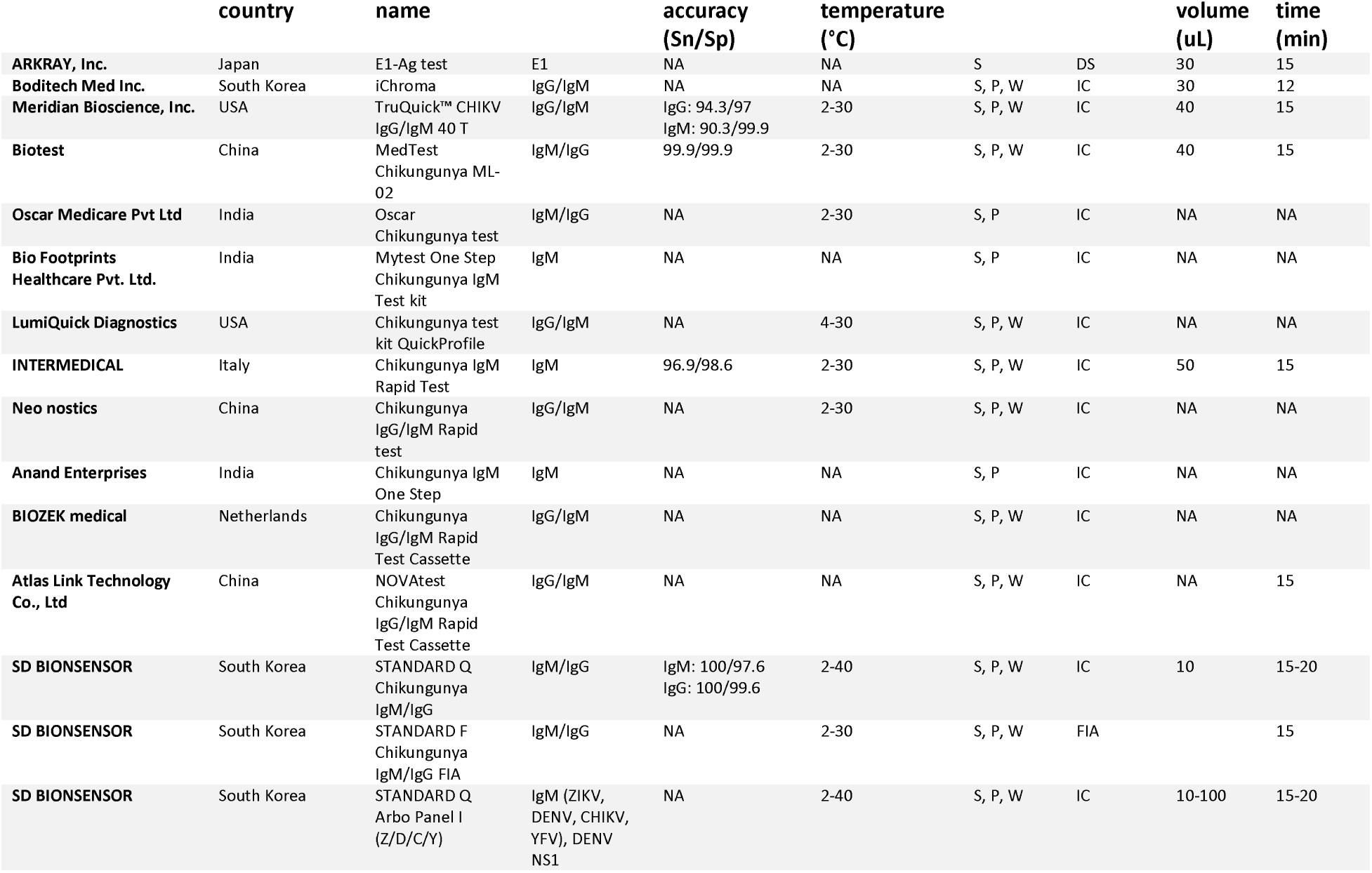
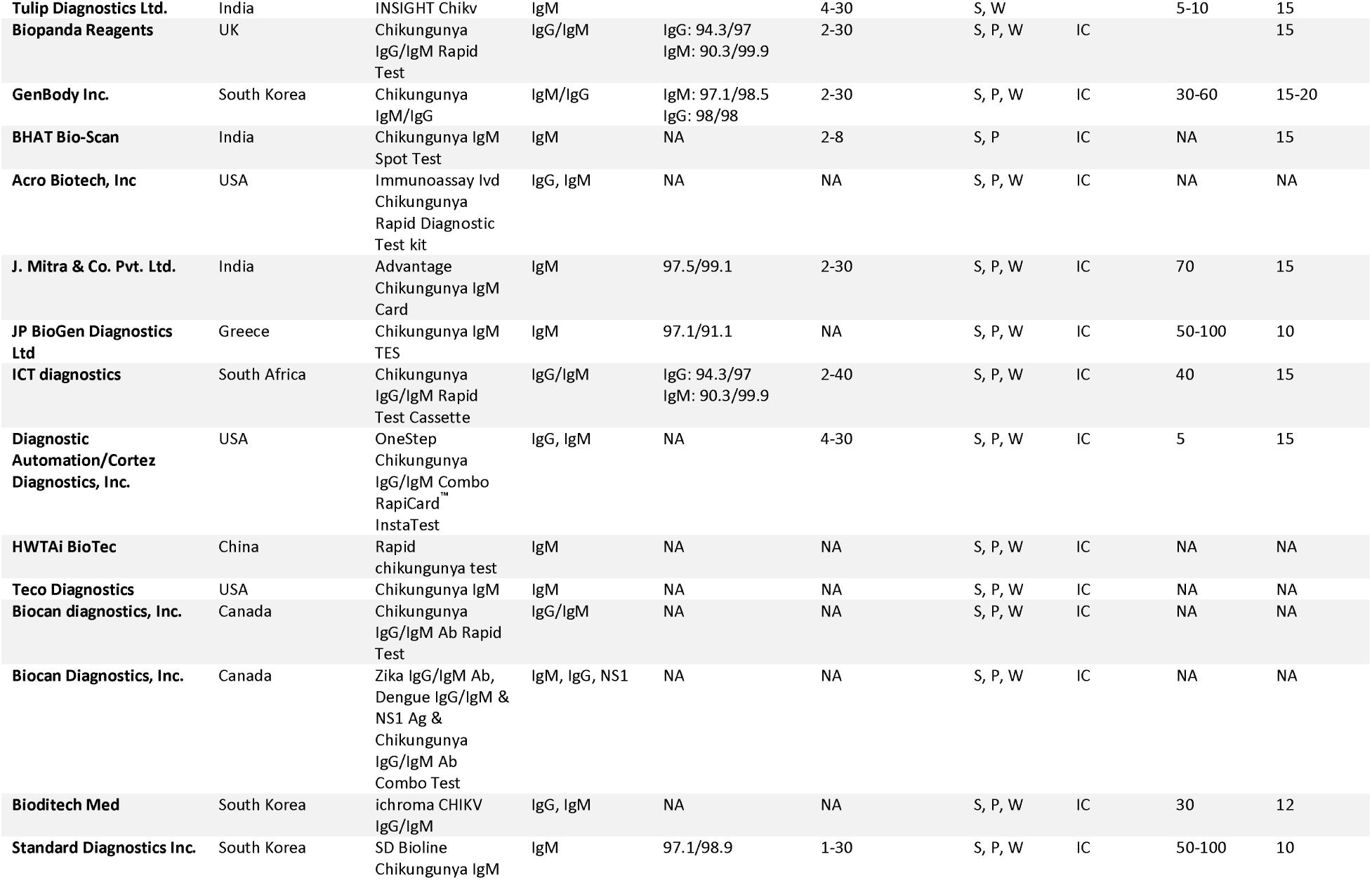
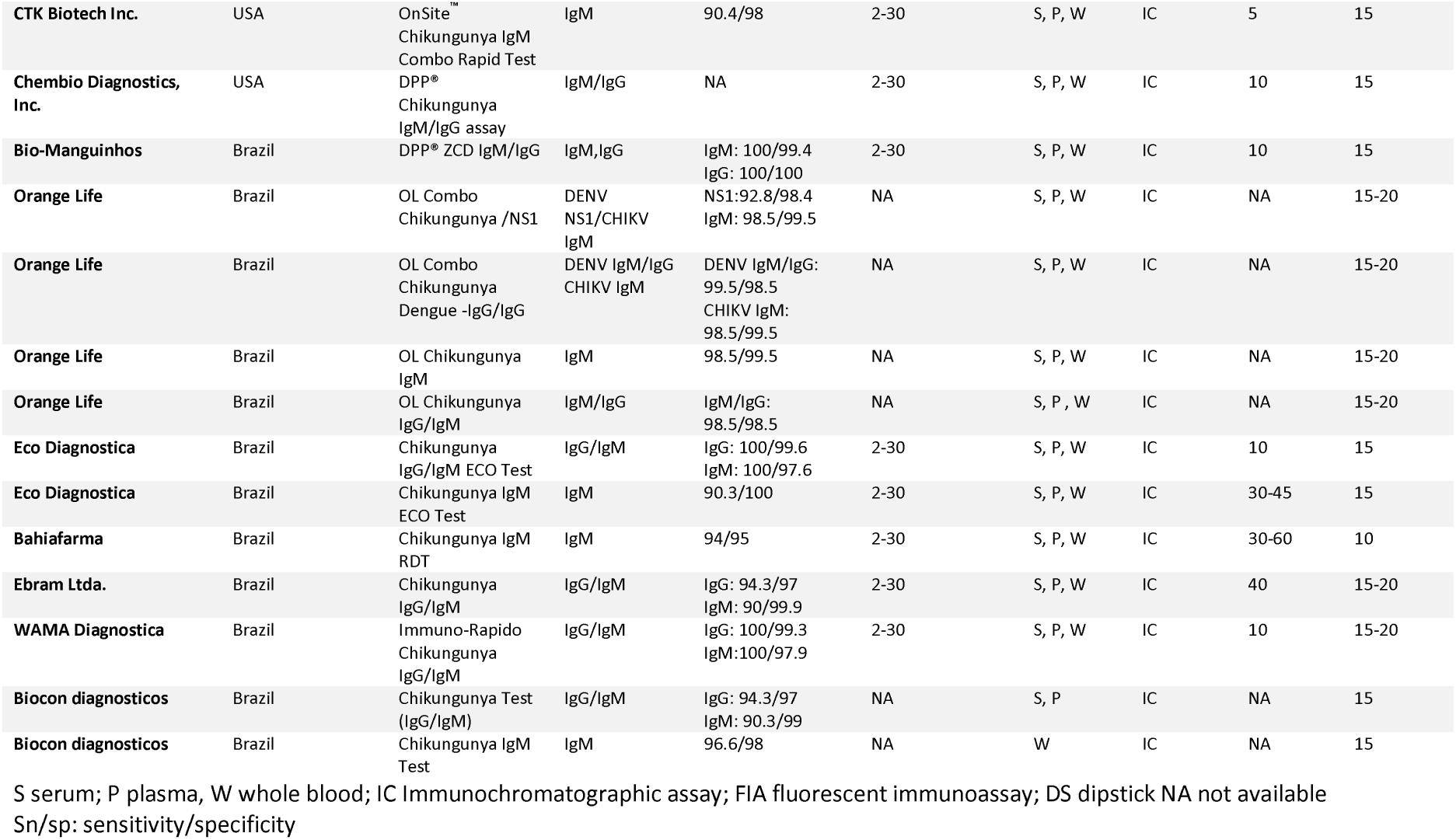
Characteristics of Chikungunya rapid diagnostic tests developed or commercialized for point-of-care application

**Figure 1.**
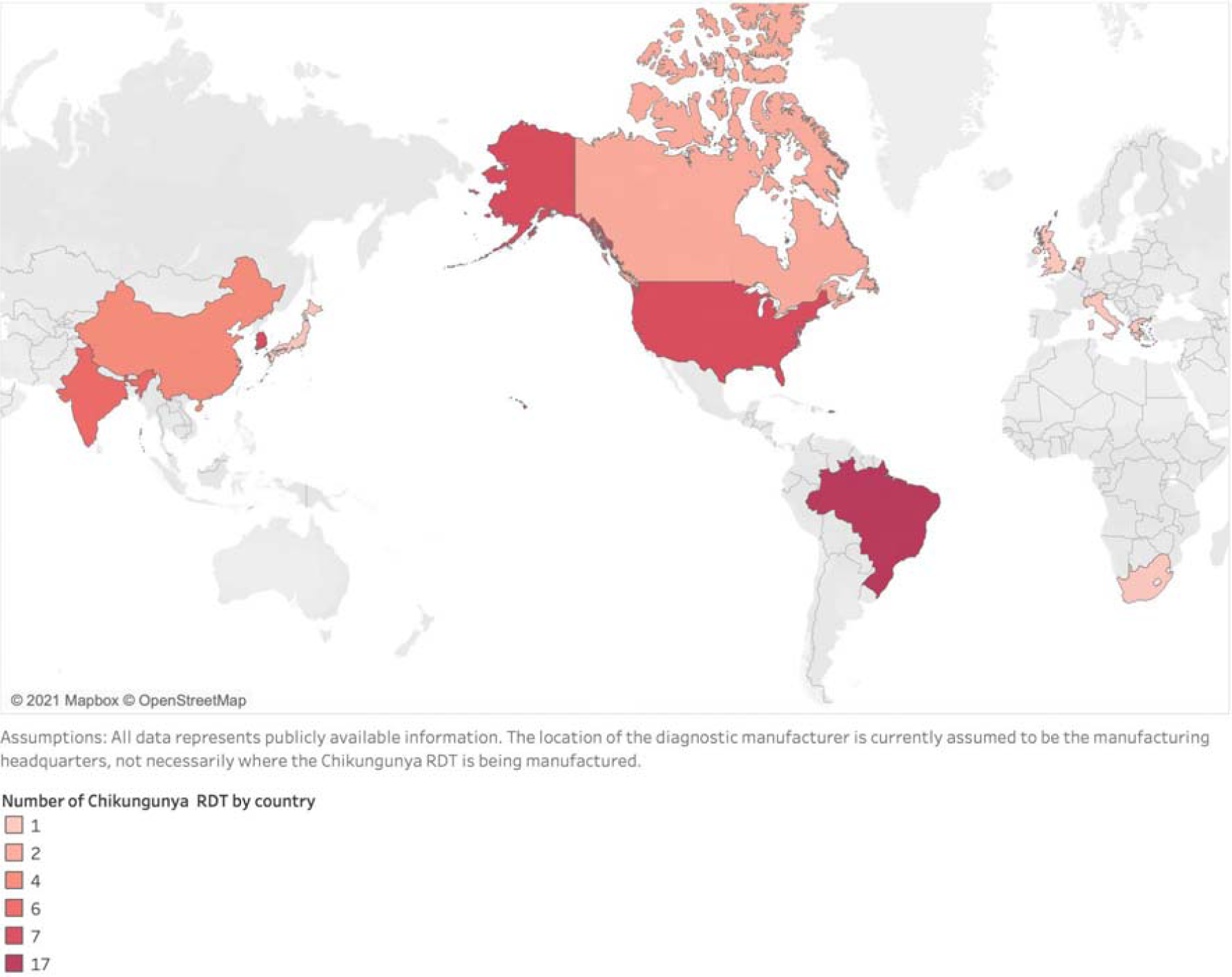
Number of Chikungunya rapid diagnostic tests developed or commercialized for point-of-care application by country of manufacture.

Overall, the CHIKV RDT market is fragmented, but the manufacturer with the most products in the market is Chembio Diagnostics Brazil (*n*= 5 products) and SD BIOSENSOR (*n*=3 products) [Table 3]. Almost all assays are antibody-based RDTs (*n*=43) designed in an immunochromatographic format. There were neither antigen-based RDTs nor a combination of antibody and antigen-based RDTs commercially available. Our searches for approved assays in national regulatory authorities did not find any assay registered by the European Medicines Agency or the U.S. Food and Drug Administration. Conversely, the Brazilian National Health Surveillance Agency (ANVISA) has approved 23 CHIKV RDTs for clinical use. Of these, 5/23 (21.7%) were multiplex assays with targets concomitant for Dengue and Zika analytes. Supplementary Table 1 shows the characteristics of CHIKV RDTs approved by the ANVISA.

**Table 3.**
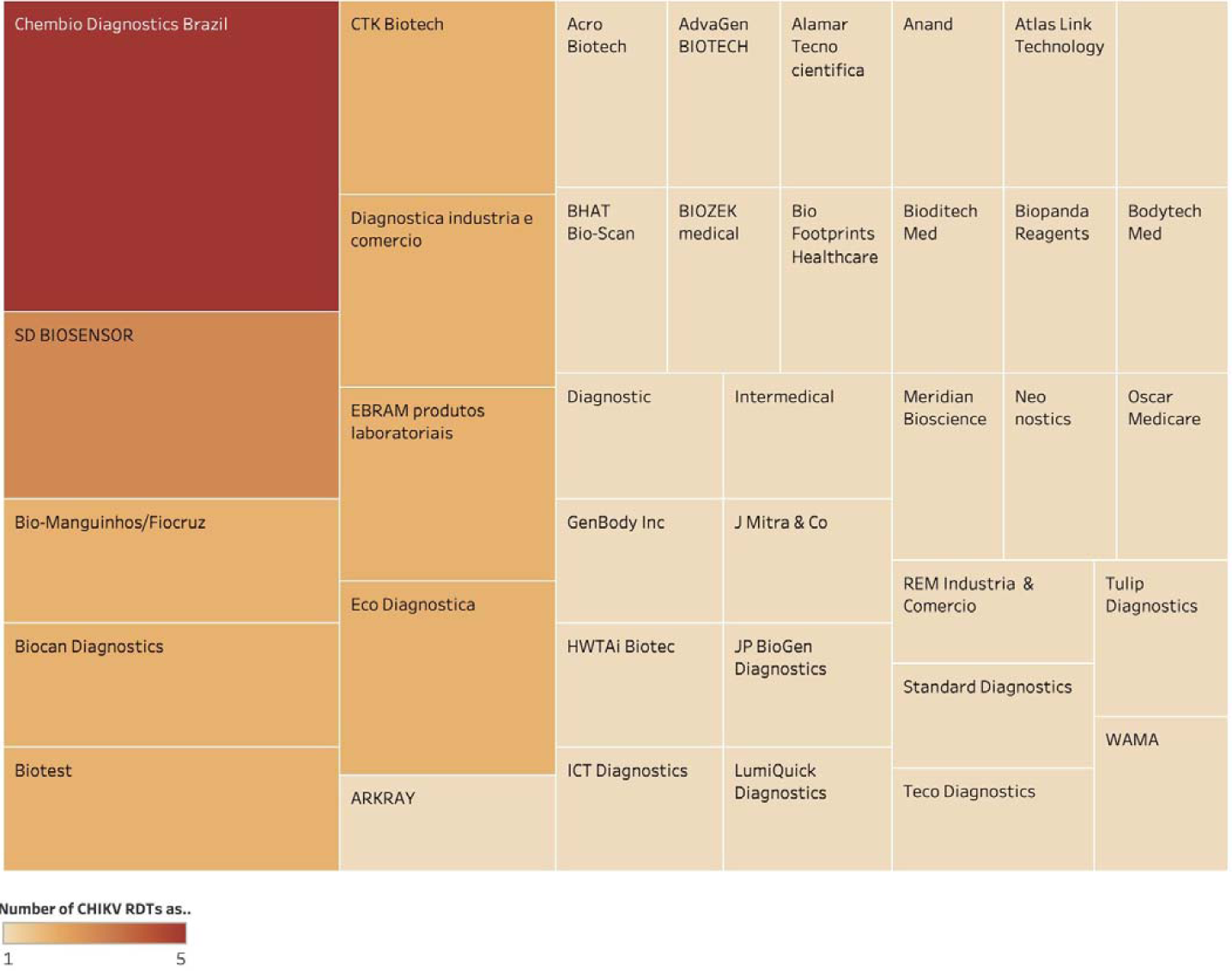
Global Chikungunya rapid diagnostic tests landscape – key players on industry

### Diagnostic accuracy results

Table 4 shows a summary of the diagnostic assessments included conducted between 2005-2019. In total, 3722 samples were tested with RDTs across all the studies (Supplementary Figure 3). Sample types included whole blood, plasma, and serum. Twelve studies examined the performance of antibody-based RDTs [11][12][13][14][15][16][17][9,18][19][9] while five the antigen-based RDTs [20][21][22][5][23].

**Table 4.**
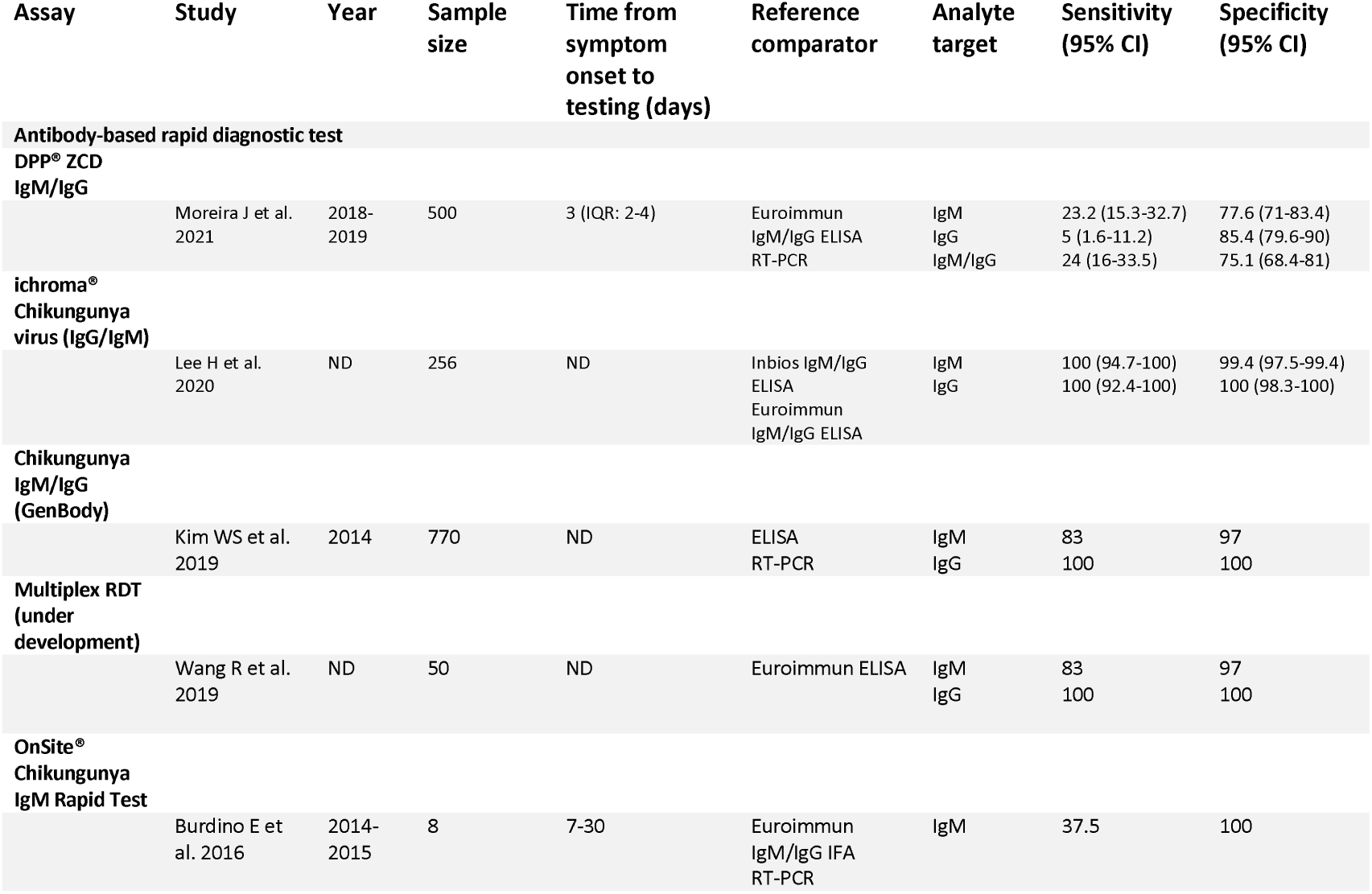
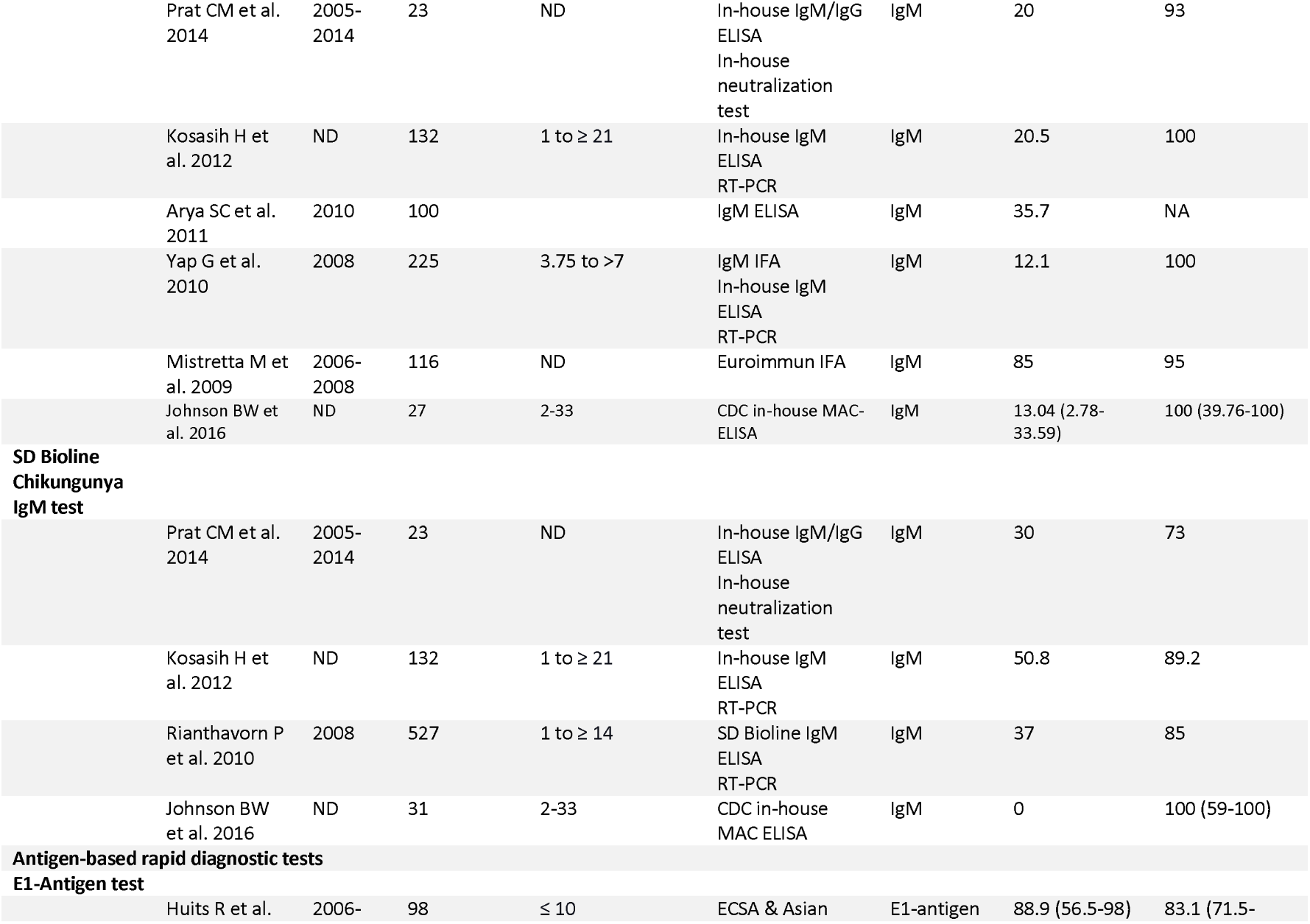
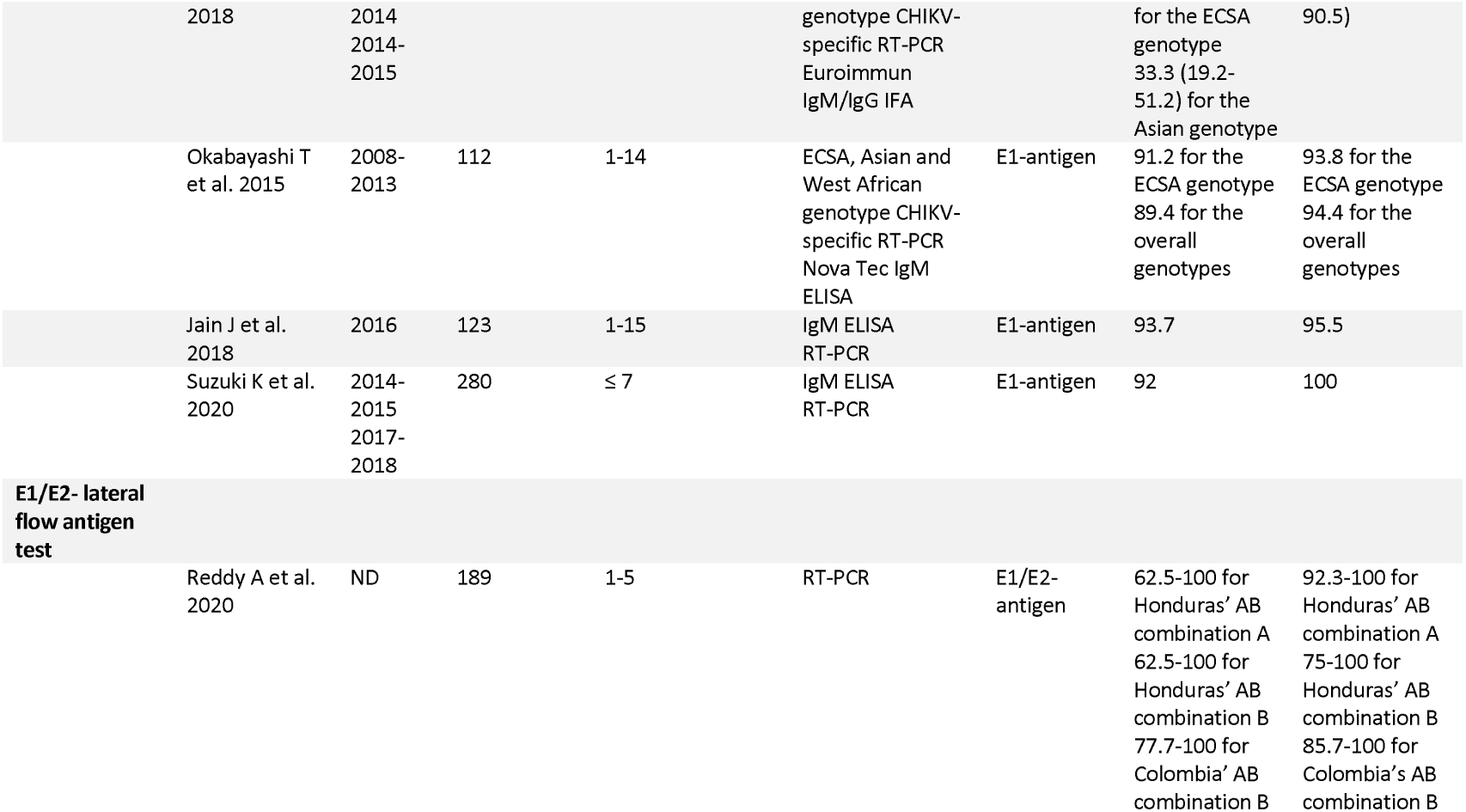
Summary of diagnostic assessments of Chikungunya antibody or antigen-based rapid diagnostic tests, 2005-2019.

The predominant CHIKV RDT assay evaluated in the studies was the OnSite® Chikungunya IgM Combo Rapid test (CTK Biotech, Inc., Poway, CA, USA) in 8/17 (47%) studies, followed by the SD BIOLINE Chikungunya IgM test (Standard Diagnostics Inc., Yongin-si, South Korea) in 3/17 (17.6%) studies. The most of antibody RDTs studies target IgM, while four studies target both IgM and IgG immunoglobulin components. Figure 2 shows the diagnostic accuracy for the OnSite® Chikungunya IgM Combo Rapid test and SD BIOLINE Chikungunya IgM test.

**Figure 2.**
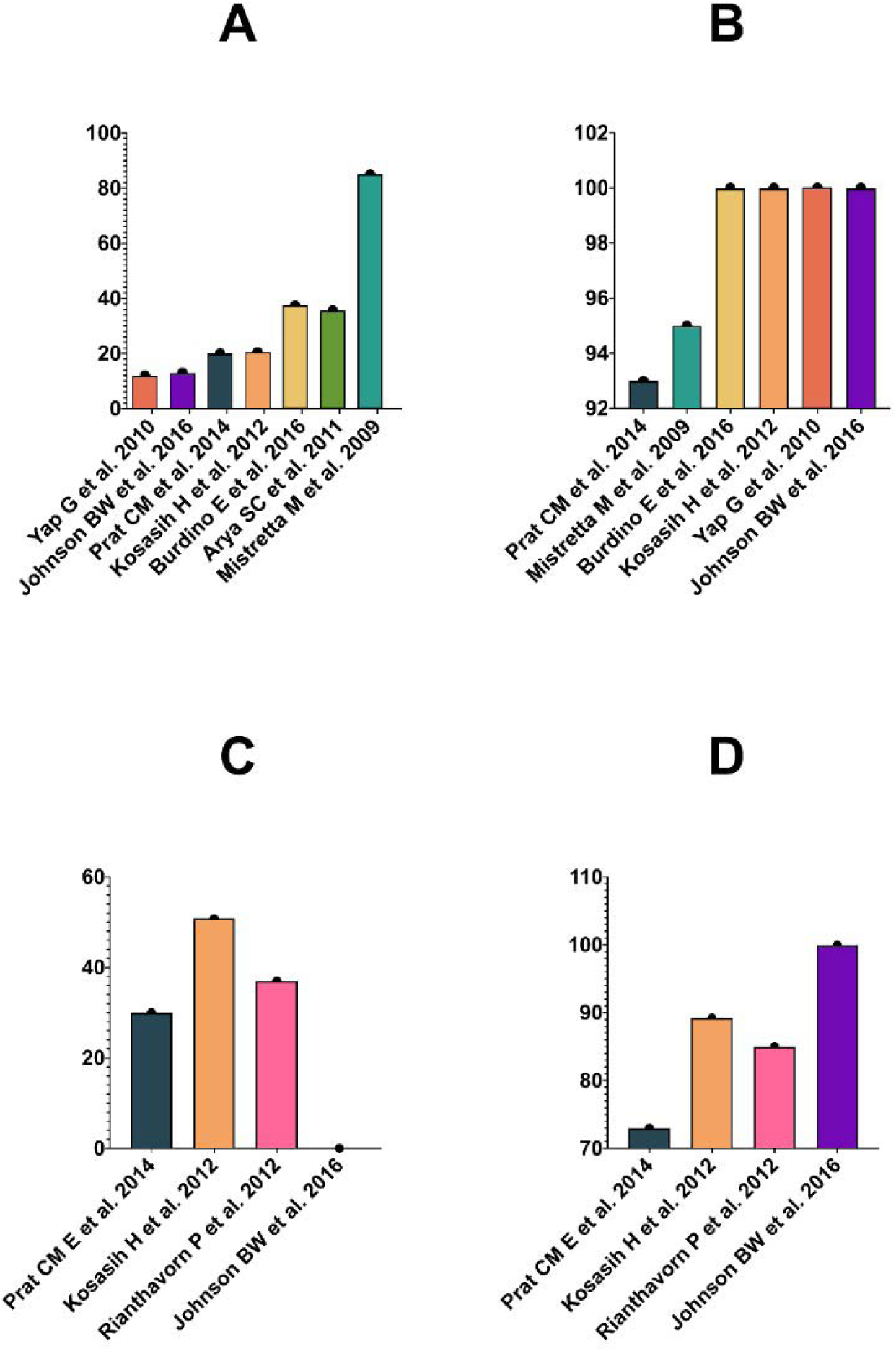
A. OnSite® Chikungunya IgM Combo Rapid test (CTK Biotech, Inc., Poway, CA, USA) sensitivity across the studies B. OnSite® Chikungunya IgM Combo Rapid test (CTK Biotech, Inc., Poway, CA, USA) specificity across the studies C. SD BIOLINE Chikungunya IgM test (Standard Diagnostics Inc., Yongin-si, South Korea) sensitivity across the studies D. SD BIOLINE Chikungunya IgM test (Standard Diagnostics Inc., Yongin-si, South Korea) specificity across the studies

Overall, the sensitivity of the RDT IgM component typically ranged between 20% and 100%. The sensitivity of the RDT IgG component ranged from 5-100%. The RDT IgM and the IgG specificity ranged from 77.6-100% and 85.4-100%, respectively. Interestingly, some studies reported an increase in the overall sensitivity of antibody-based RDT over time [[4][15]. There are two types of antigen-based RDTs evaluated – E1 and E1/E2-antigens tests. The sensitivity of the E1-antigen tests ranged from 33.3 to 100%. Conversely, the specificity varied between 83.1-100%.

### Risk of bias assessment

Figure 3 summarizes the QUADAS-2 assessment by study. There were patient selection applicability concerns for most of the study (*n*=14) because there was a lack of sufficient information reported in the studies regarding the patient population, demographic features, setting of the study, or presence of co-morbidities. Similarly, there was a high risk of bias in the patient selection domain because only three studies enrolled a consecutive or random sample of eligible patients with suspicion of CHIKV infection to reduce the bias in the diagnostic accuracy of the index test.

**Figure 3.**
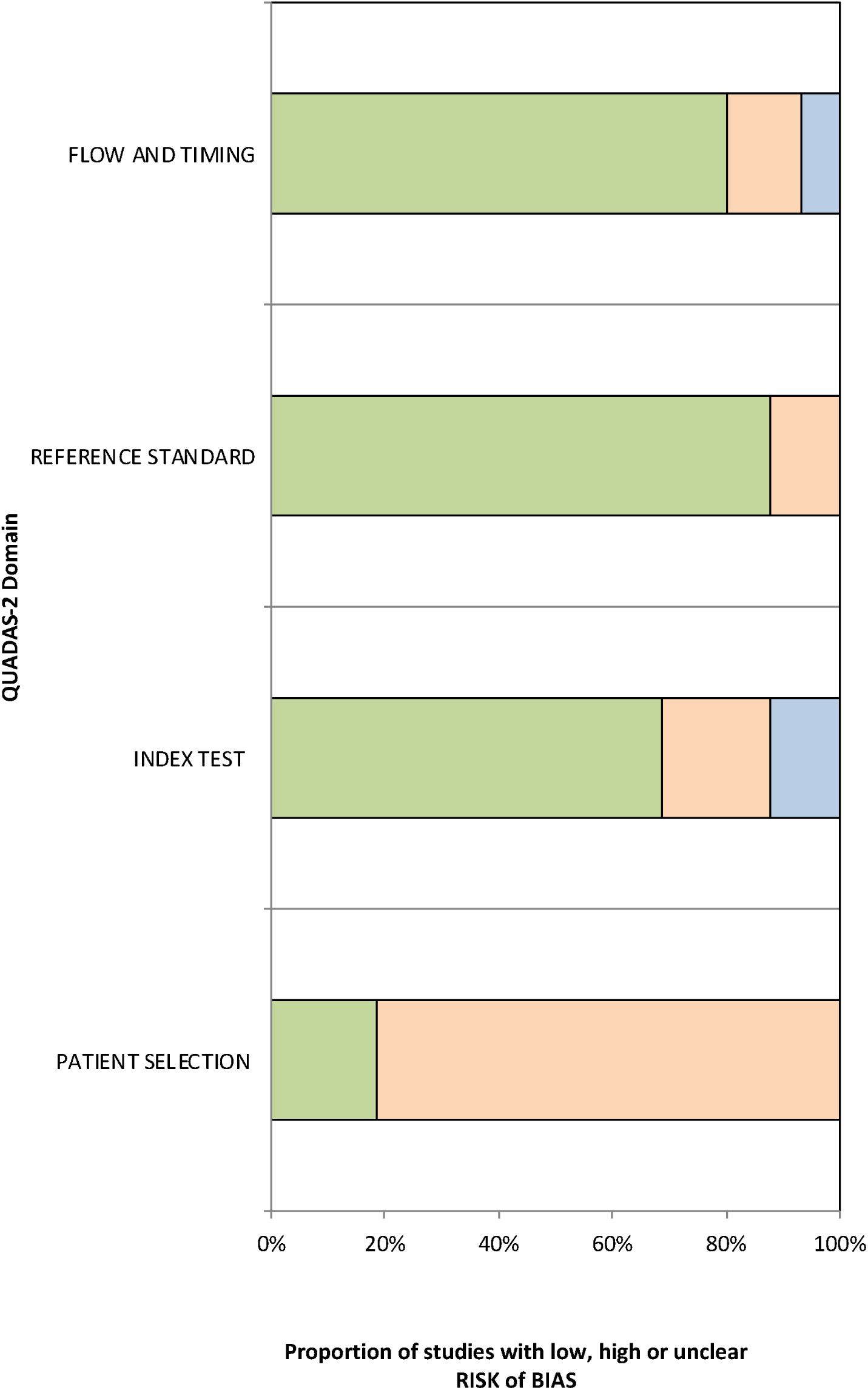
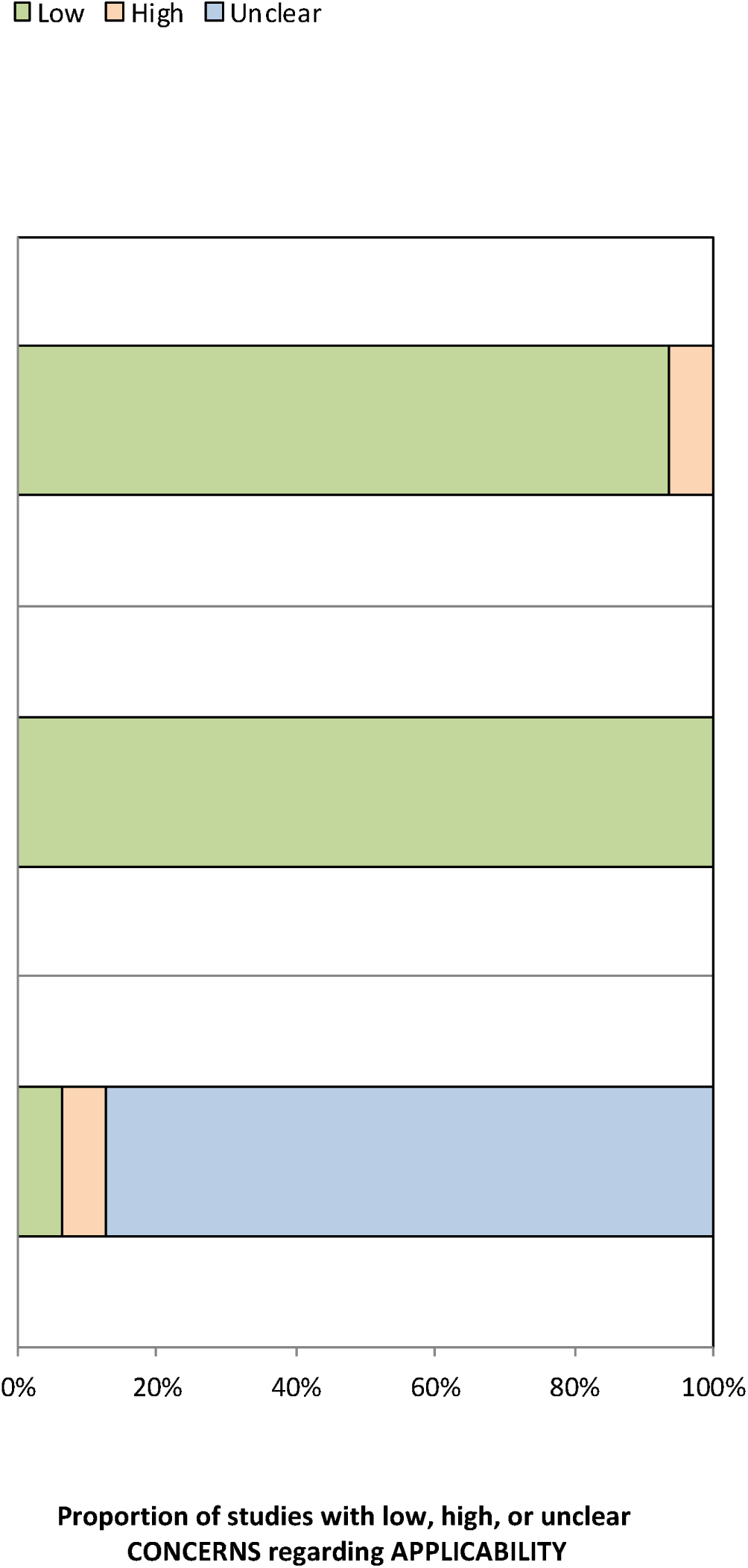

## Discussion

### Summary of evidence

This scoping review identified 18 studies conducted between 2005 and 2019, addressing the research stage on CHIKV RDTs across various settings. Our findings indicate a paucity of research focusing on field trials and implementation studies related to CHIKV RDTs. Our work provides a global view of publicly available data on CHIKV RDTs currently under development or commercially available. We also found that the in vitro diagnostic medical device manufacturers are primarily concentrated on CHIKV antibody RDTs, and their accuracy overall performs poorly and should not be used in clinical settings as long as they suffer significant improvements [15][4]. Conversely, antigen RDTs, although still in a development phase, promise to have a high level of sensitivity and specificity across the distinct CHIKV genotypes [5][22].

Given the problems associated with the existing diagnostic strategies for CHIKV, there is a clear and urgent need for new, appropriate diagnostic tools for CHIKV that meet the ideal product profile of “REASSURED” diagnostics [24]. The characteristics of the diagnostics products mentioned above are defined by a set of criteria which includes: (i) Real-time connectivity; (ii) Ease of specimen collection; (iii) Environmental friendliness; (iv) Affordable by those at risk of infection; (v) Sensitive (few false-negatives); (vi) Specific (few false-positives); (vii) User-friendly (simple to perform and requiring minimal training); (viii) Rapid (to enable treatment at first visit) and Robust (does not require refrigerated storage); (xi) Equipment-free; and (x) Delivered to those who need it. Few products right now meet the ideal “REASSURED” profile, and new research and investments are required to develop those that match the profile needed. Pertinent questions about feasibility, acceptability, cost-effectiveness, sustainability, and policy implications must be addressed before the widespread use of CHIKV RDTs in endemic countries. More importantly, we also need to address the impact of CHIKV RDTs into integrated fever case management and how its implementation translates into a better prescription practice for acute febrile patients (i.e., reducing unnecessary antibiotic prescription).

The CHIKV RDTs diagnostic landscape is fragmented, with many gaps along the development pathway. Figure 4 shows our proposed conceptual framework that delineates the challenges and opportunities across each stage of CHIKV RDT development. Concerted efforts leading by different stakeholders (i.e., international donors, industry, public sector, and end-users) should be put together to bring more equity to the availability of appropriate CHIKV RDTs to those needed most.

**Figure 4.**
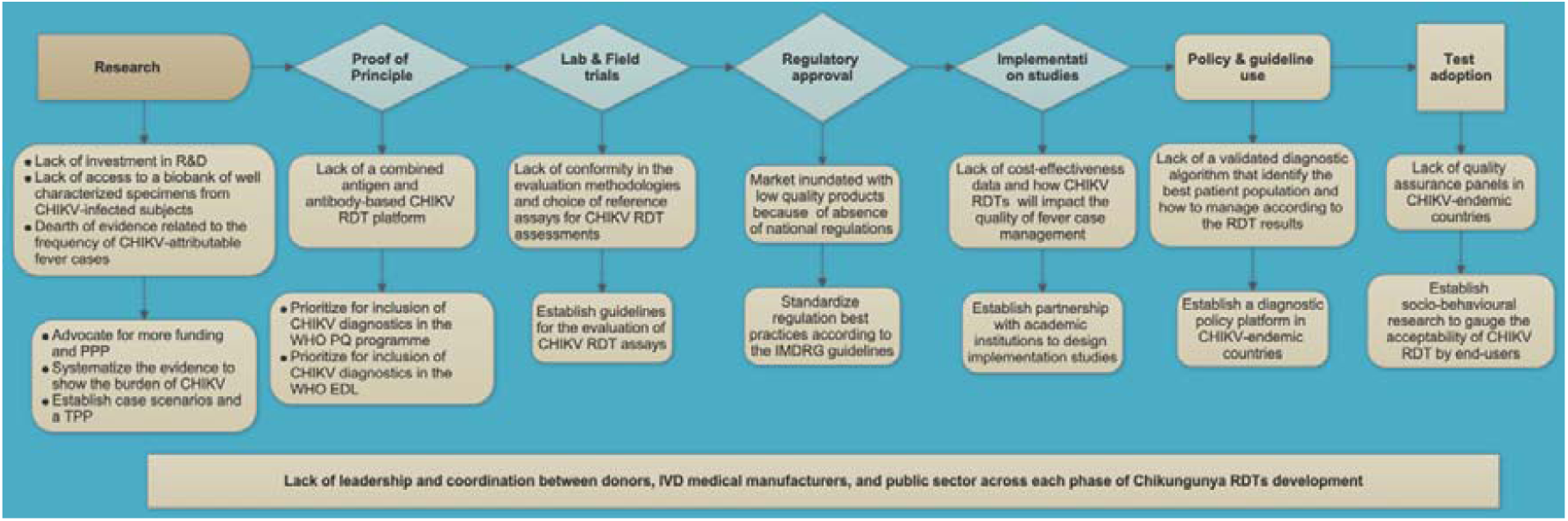
Chikungunya rapid diagnostic tests: Fragmented landscape presents market challenges and opportunities for interventions CHIKV chikungunya virus; RDT rapid diagnostic test; EDL World Health Organization’ Essential diagnostic list; IMDRG International Medical Devices Regulatory Forum; IVD in-vitro diagnostic products; PPP Public-Private Partnership; P.Q. World Health Organization’ in-vitro diagnostic pre-qualification program; R&D Research & Development; TPP target product profile; WHO World Health Organization

### Limitations

Our work has limitations. Although we made a herculean effort to identify the highest numbers of CHIKV RDTs manufactured or commercially available in the market, we understand that some could not be identified and were not publicly available. However, we addressed this bias by looking into CHIKV RDTs that national/regional regulatory agencies have approved or those that provided data from unpublished sources (i.e., conference abstracts, manufacturers’ reports, author’s unpublished data). Next, we did not provide an effect estimate for the results of diagnostic accuracy studies, because as shown in our risk of bias assessment, the studies included were very heterogeneous, and a meta-analytic approach would be useless.

### Conclusions

Our scoping review demonstrated substantial gaps in the current diagnostic landscape of CHIKV RDTs. The future needs of immunoassay-based RDTs for CHIKV are summarized in Box.

The time is suitable for a collaborative, focused initiative between policy-makers and other relevant stakeholders to address the urgent need for new, appropriate CHIKV RDTs. Unprecedented opportunities for market interventions exist and utilize new technologies to make a significant, measurable impact. Further research is desperately needed to facilitate the incorporation of CHIKV RDTs into integrated fever algorithms, and socio-behavioral research should be done to evaluate end-user acceptability.

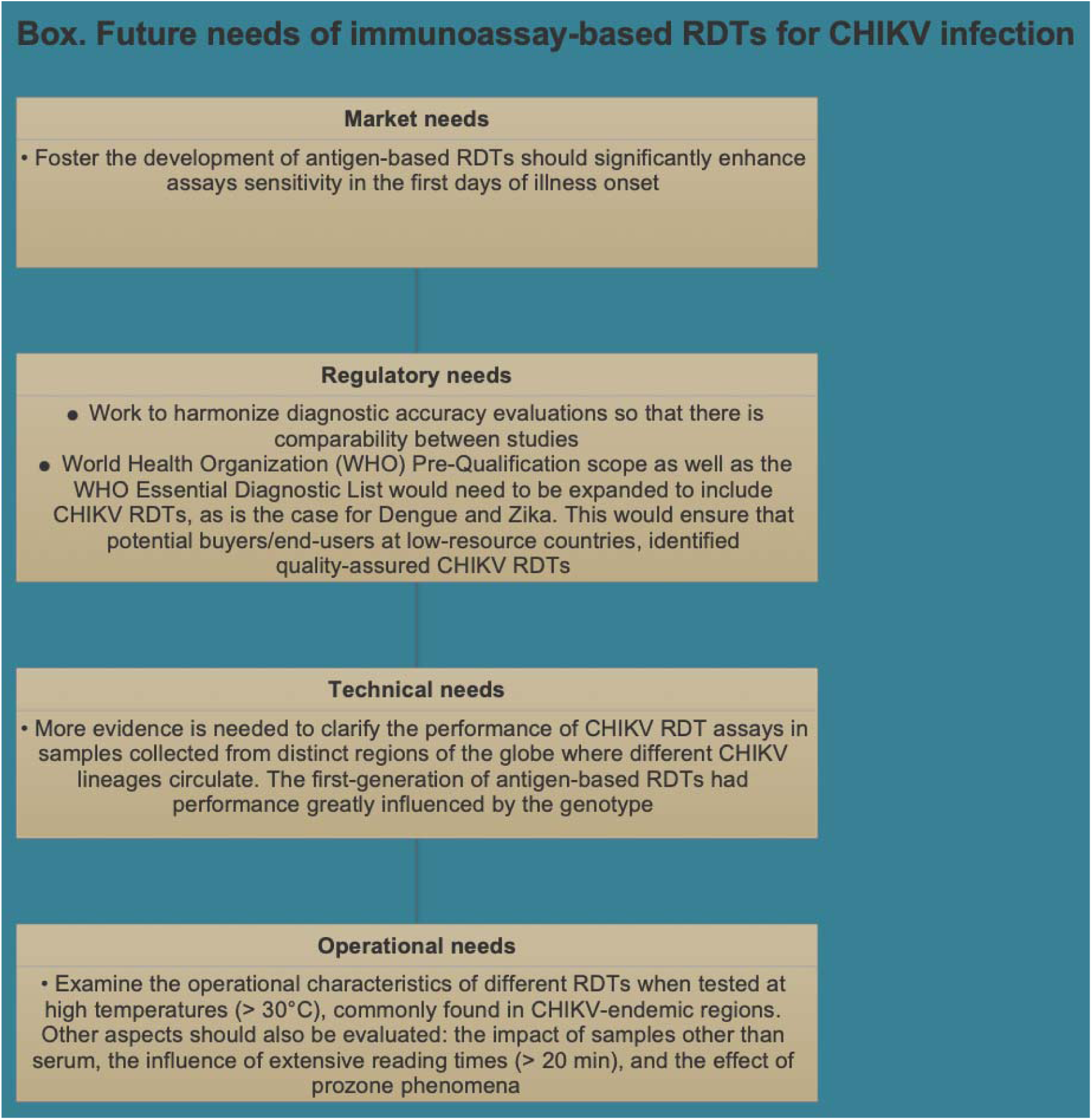

## Data Availability

All data produced in the present work are contained in the manuscript.

## Contributions

All authors critically reviewed the manuscript, gave their final approvals, and are accountable for accuracy and integrity.

## Acknowledgments

J.M. received a scholarship from Fundacao de Amparo a Pesquisa do Estado do Rio de Janeiro (FAPERJ) for his Ph.D. study in Brazil.

## Figure legend

Figure 1. Number of Chikungunya rapid diagnostic tests developed or commercialized for point-of-care application by country of manufacture

Figure 2. Summary of diagnostic accuracy studies evaluating the OnSite® Chikungunya IgM Combo Rapid test (CTK Biotech, Inc., Poway, CA, USA) and the SD BIOLINE Chikungunya IgM test (Standard Diagnostics Inc., Yongin-si, South Korea)

Figure 3. QUADAS-2 assessment of studies

Figure 4. Chikungunya rapid diagnostic tests: Fragmented landscape presents market challenges and opportunities for interventions

