## Supplementary Figures & Tables for "Mapping The Global Landscape Of Chikungunya Rapid Diagnostic Tests: A Scoping Review"

**Appendix Table of Contents**

This supplementary material has been provided by the authors to give readers additional information about their work.

**Supplementary Table**

- Supplementary Table 1. Characteristics of commercial Chikungunya rapid diagnostic tests for point-of-care application registered by the Brazilian National Health Surveillance Agency

**Supplementary Figures**

- Supplementary Figure 1. PRISMA flowchart diagram
- Supplementary Figure 2. Sources of Chikungunya samples evaluated for rapid diagnostic test, 2005-2019
- Supplementary Figure 3. Number of samples tested according to Chikungunya rapid diagnostic test, 2005-2019

Supplementary Figure 1. PRISMA flowchart diagram

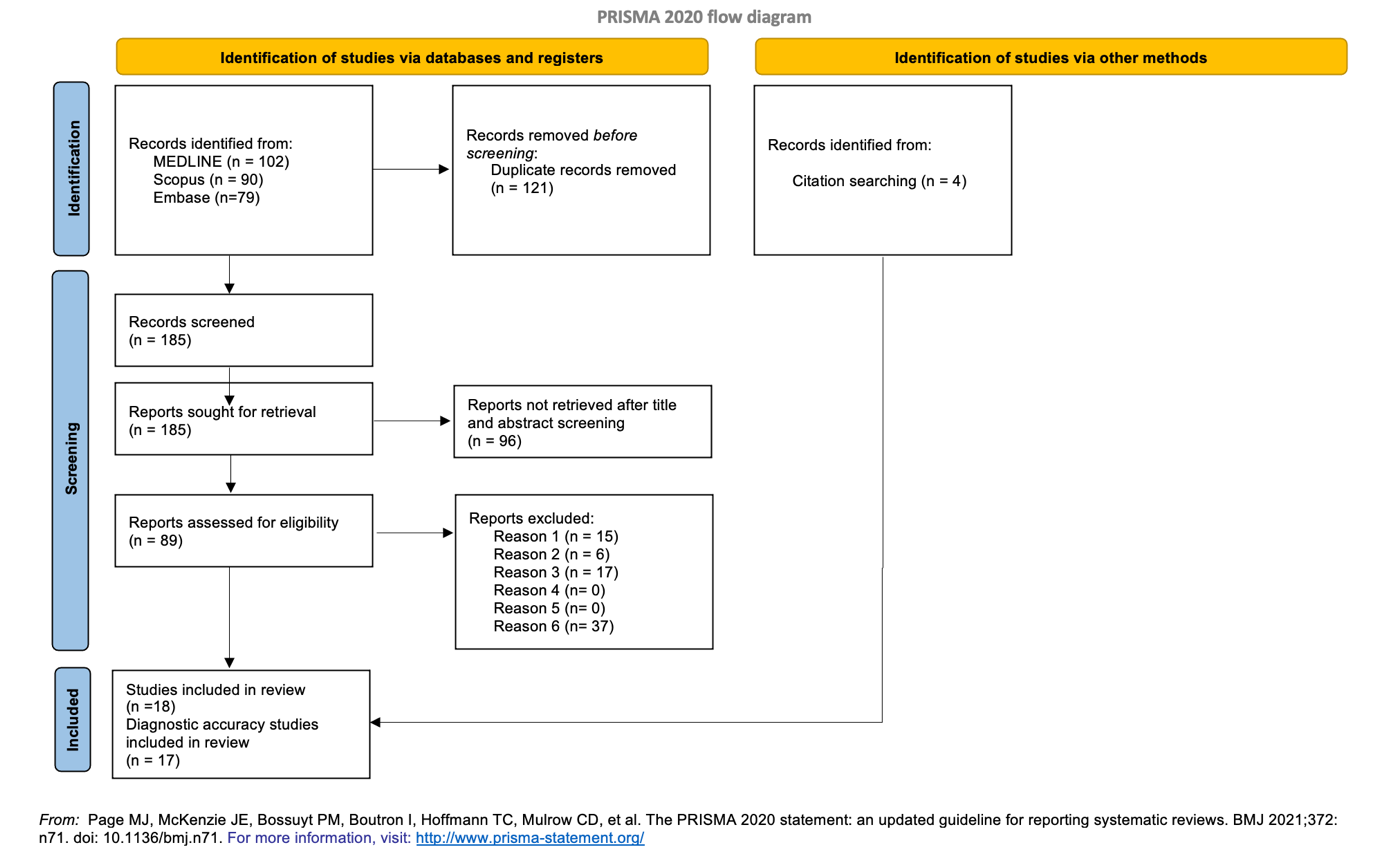

Supplementary Figure 2. Sources of Chikungunya samples evaluated for rapid diagnostic test, 2005-2019

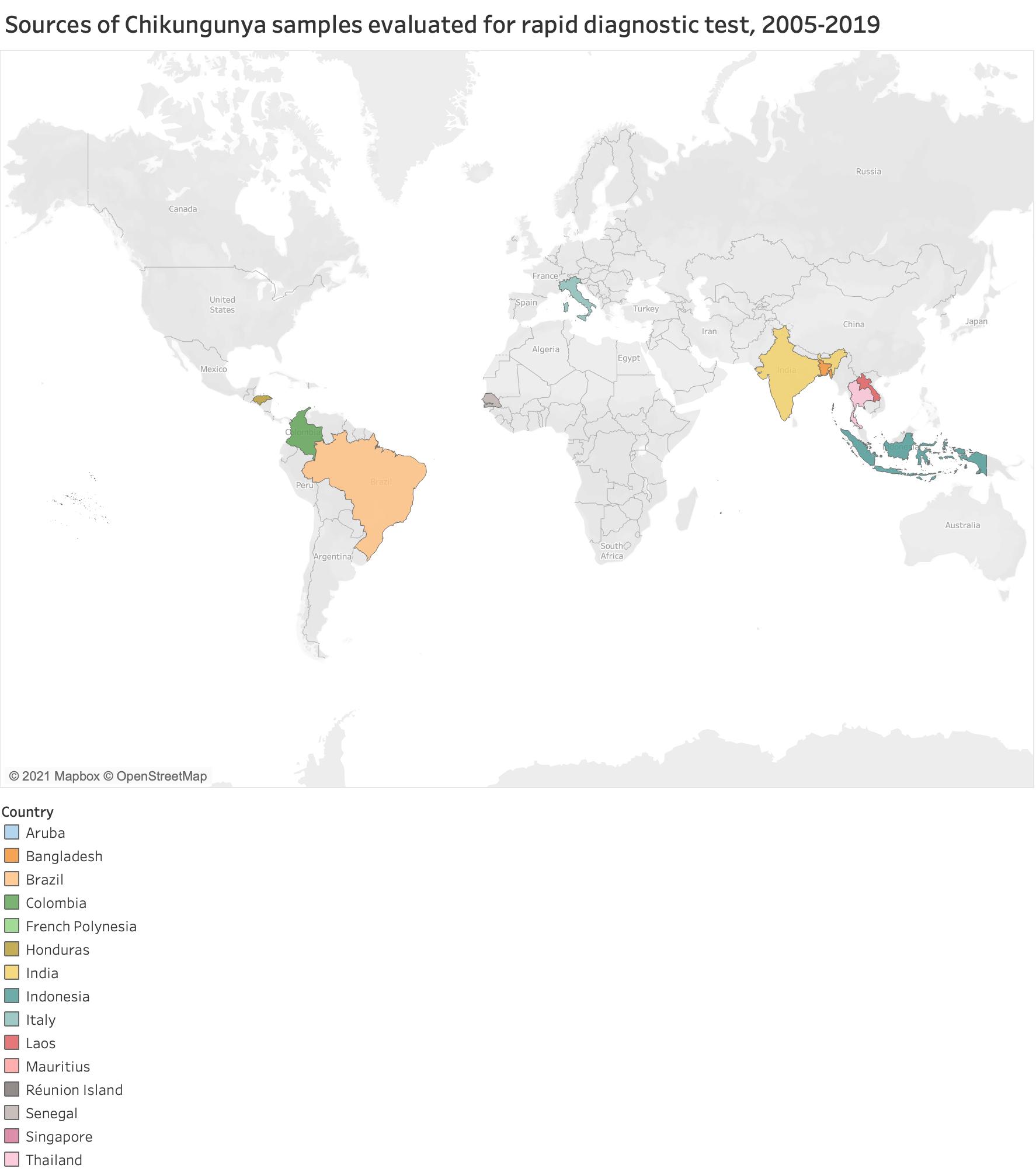

Supplementary Figure 3. Number of samples tested according to Chikungunya rapid diagnostic test, 2005-2019

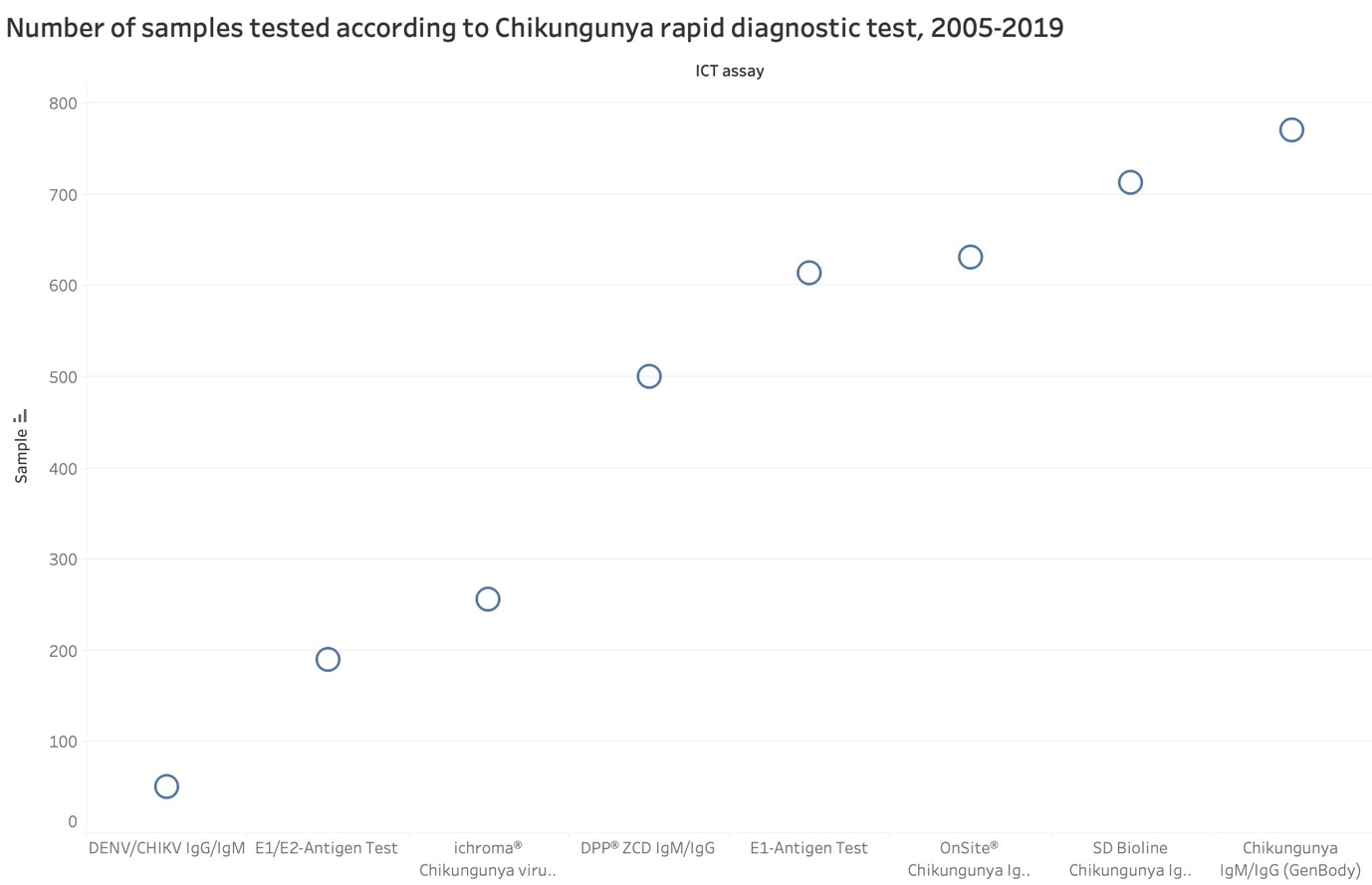

ICT stands for immunochromatography assay

Supplementary Table 1. Characteristics of commercial Chikungunya rapid diagnostic tests for point-of-care application registered by the Brazilian National Health Surveillance Agency.

| Product name | Manufacturer or importer | Manufacture country | Package presentation | Multiplex test (target) | Registration no. | Registration due date |
| --- | --- | --- | --- | --- | --- | --- |
| Chikungunya LF IgG IgM | AdvaGen BIOTECH LTDA | Brazil | 10, 20, 25, 50, or 100 tests | No | 81472060015 | 23-09-29 |
| Chikungunya IgG/IgM test rapido | Alamar Tecno cientifica Ltda | Brazil | 20,25, 50, or 100 tests | No | 80049120107 | 14-08-27 |
| Teste Rapido OnSite Duo Dengue IgG/IgM-CHIKV IgM | CTK Biotech, Inc.. | USA | 30 tests | Yes (DENV) | 80524900030 | 30-11-25 |
| Teste Rapido OnSite Chikungunya IgM | CTK Biotech, Inc. | USA | 30 tests | No | 80524900031 | 30-11-25 |
| OL Chikungunya IgM | Chembio Diagnostics Brazil Ltda. | Brazil | 1, 5, 10, 20, 25, 30, 50, or 100 tests | No | 80535240028 | 22-09-24 |
| OL Combo Chikungunya – Dengue IgG/IgM | Chembio Diagnostics Brazil Ltda. | Brazil | 1, 5, 10, 20, 25, 30, 50, or 100 tests | Yes (DENV) | 80535240034 | 05-01-25 |
| OL Combo Chikungunya/NS1 | Chembio Diagnostics Brazil Ltda. | Brazil | 1, 5, 10, 20, 25, 30, 50, or 100 tests | Yes (DENV) | 80535240036 | 02-02-25 |
| OL Combo Chikungunya-Dengue NS1/IgG/IgM | Chembio Diagnostics Brazil Ltda. | Brazil | 1, 5, 10, 20, 25, 30, 50, or 100 tests | Yes (DENV) | 80535240037 | 02-02-25 |
| OL Chikungunya IgG/IgM | Chembio Diagnostics Brazil Ltda. | Brazil | 1, 5, 10, 20, 25, 30, 50, or 100 tests | No | 80535240038 | 30-03-25 |
| Chikungunya IgG/IgM Rapid Test | Diagnostica indústria e comercio Ltda | Brazil | 10, 20 or 25 tests | No | 80638720070 | 12-12-26 |
| Chikungunya IgM Rapid Test | Diagnostica indústria e comercio Ltda | Brazil | 10, 20, 30, 40, 60, 70, 80, 90, or 100 tests | No | 80638720107 | 27-05-29 |
| Chikungunya IgM | EBRAM produtos Laboratoriais Ltda | Brazil | 10, 20, 25, or 50 tests | No | 10159820208 | Expired in 16-09-19 |
| Chikungunya IgG/IgM | EBRAM produtos Laboratoriais Ltda | Brazil | 10, 40, or 100 tests | No | 10159820231 | 25-11-29 |
| CHIKV IgG/IgM ECO Teste | Eco Diagnostica Ltda | Brazil | 1, 10, 25, 30, 50, or 100 | No | 80954880023 | 02-05-27 |
| ECO F CHIKV IgG/IgM | Eco Diagnostica Ltda | Brazil | 1, 5, 10, 20, 25, 30, 40, 50 or 100 tests | No | 80954880069 | 14-05-28 |
| Teste Rapido Chikungunya IgM Bahiafarma | Fundacao Baiana de Pesquisa Cientifica e Desenvolvimento Tecnologico, Fornecimento e Distribuicao de Medicamentos-BAHIAFARMA | Brazil | 20,50, or 100 tests | No | 81285200002 | 05-09-26 |
| TR DPP® CHIKUNGUNYA IgM/IgG – Bio-Manguinhos | Fundacao Oswaldo Cruz | Brazil | 20 tests | No | 80142170034 | 06-08-28 |
| TR DPP® ZDC IgM/IgG – Bio-Manguinhos | Fundacao Oswaldo Cruz | Brazil | 20 tests | Yes (ZIKV & DENV) | 80142170035 | 08-04-29 |
| MedTeste Chikungunya | Hangzhou Biotest Biotech Co. Ltd. | China | 1, 10, 20, 25, 50 or 100 tests | No | 80560310046 | 06-05-29 |
| MedTeste Chikungunya ML-02 (Teste Rapido) | Hangzhou Biotest Biotech Co. Ltd. | China | 1, 10, 20, 25, 50 or 100 tests | No | 80560310052 | 17-02-30 |
| QuickProfile® Chikungunya IgG/IgM Combo Test Card | Lumiquick Diagnostics, Inc | USA | 25 | No | 81268670001 | 30-01-27 |
| Gold Rapid Test Chikungunya IgG/IgM | REM Industria e Comercio Ltda | Brazil | 25 | No | 81077790012 | 12-03-28 |
| Imuno-Rapido Chikungunya IgG/IgM | WAMA | China | 10, 20, 25, 40, or 80 tests | No | 10310030182 | 01-03-27 |

DENV stands for Dengue virus; ZIKV Zika virus; USA United States of America
